# An investigation of dental-related emergency department presentations and patient characteristics in Victoria, Australia

**DOI:** 10.64898/2026.09.24.26363871

**Authors:** Radiya Adam, Jessica Nguyen, Austen Keating, Clare Lin, Tan Minh Nguyen

## Abstract

**Objective:** Describe the profile of consumers utilising Emergency Departments (ED) across Victorian hospitals for dental conditions between financial year 2018-19 to 2022-23.

**Methods:** Dental conditions were grouped according to the diagnosis categories using the International Statistical Classification of Diseases and Related Health Problems, Tenth Revision, Australian Modification (ICD-10-AM). Presentations for non-dental conditions were classified as “Other”. Dependent variables included age, sex, Aboriginal and Torres Strait Islander (Aboriginal) background, language other than English, interpreter required, remoteness, and socio-economic disadvantage according to the Index of Relative Socio-economic Disadvantage (IRSD).

The primary outcome evaluated is the relative risk ratio (RRR) of having a hospital ED presentation for a dental condition compared to a non-dental condition and age-specific rates per 100,000 population. Multinomial logistic regression was performed following the backwards elimination method.

**Results:** Over the years 2018-19 to 2022-23, 0.55% (n=44,382) of ED presentations across Victorian hospitals by Victorian residents were due to dental conditions. Patients aged 20-44 years have the highest RRR compared to other age groups for dental-related ED presentations. Patients experiencing social inequity in terms of demographic factors such as Aboriginal background, requiring an interpreter, not living in a major city and living in areas of greater disadvantage (IRSD) have statistically significant increased RRR for dental-related ED presentations.

**Conclusion:** Patient characteristics were useful for identifying the relative risk of dental-related ED presentations between socio-economic and demographic groups to inform relevant public policy design focus on preventing dental-related ED.

## Introduction

Inequities in oral health are caused by a complex array of individual, social, environmental, economic and political determinants (de Abreu et al., 2021; Tsakos et al., 2023). Socio-economic position influences oral healthcare utilisation, oral hygiene and free sugar consumption, disproportionately increasing the risk of poorer oral health in people with low socio-economic status (de Abreu et al., 2021; Tsakos et al., 2023). When oral disease is not treated in a timely manner through effective prevention and primary care, people may have no other choice than to visit the emergency department (ED) for disease management.

Dental-related ED presentations are uncommon but costly events across major hospitals in Australia. Studies have shown that these cases accounted for approximately 1% of all ED presentations in South Australia (SA) and Tasmania (Musa et al., 2025; Verma & Chambers, 2014). A single dental-related ED presentation is costly to the healthcare system with recent estimates pricing each presentation at approximately AUD$619 (Nous Group, 2025).

There are no studies that explore associations between patient characteristics and dental-related ED presentations in Victoria, Australia. To address this gap, this study aims to describe the profile of consumers utilising ED for dental conditions in Victorian between financial year 2018-19 to 2022-23.

## Methods

This study received ethics approval by Deakin University Human Research Ethics Committee (DUHREC 2024-054) and reported following the Standards for Quality Improvement Reporting Excellence (SQUIRE) guidelines (Ogrinc et al., 2016). Non-identifiable data was requested and provided by the Victorian Department of Health using the Victorian Emergency Minimum Dataset, which also included the Victorian Virtual Emergency Department. The data excluded visit by type for Coronavirus disease 2019 (Covid-19) screening patients.

Presentations for non-dental conditions were classified as “Other”. Dental conditions were grouped according to the principle diagnosis categories using the International Statistical Classification of Diseases and Related Health Problems, Tenth Revision, Australian Modification (ICD-10-AM) (Independent Health and Aged Care Pricing Authority, 2022):

- K02-Dental caries
- K03-Other diseases of hard tissues of teeth
- K04-Diseases of pulp and periapical tissues
- K05-Gingivitis and periodontal diseases
- K06-Other disorders of gingiva and edentulous alveolar ridge
- K07-Dentofacial anomalies
- K08-Other disorders of teeth and supporting structures
- K09-Cysts of oral region, not elsewhere classified
- K10-Other diseases of jaws
- K11-Diseases of salivary glands
- K12-Stomatitis and related lesions

Dependent variables included age groups (0-4, 5-9, 10-19, 20-44, 45-64, 65+ years), sex (male, intersex, female), Aboriginal and Torres Strait Islander background (Aboriginal; yes, no), language other than English (LOTE; yes, no), interpreter required (yes, no), remoteness by geographical areas defined by the Australian Statistical Geography Standard Edition 3 Remoteness Areas for Australia, (excluding Very Remote for Victoria; Australian Bureau of Statistics, Jul2021-Jun2026), usual accommodation, mental health status and quintile level of socio-economic disadvantage according to the Index of Relative Socio-economic Disadvantage (IRSD) (Australian Bureau of Statistics, 2021).

The primary outcome evaluated is the relative risk ratio (RRR) of having a dental-related ED presentation compared to a non-dental related presentation. Multinomial logistic regression was performed following the backwards elimination method (Chowdhury & Turin, 2020). This process involves including all dependent variables potentially associated with the primary outcome in the first model fit, before refitting the model with deleted variables parameters with the highest p-value, until the remaining variables is significant at the cut-off p-value, using the conventional threshold of 0.05 (Chowdhury & Turin, 2020).

Crude age-specific rates per 100,000 population were calculated in Excel to understand dental-related ED presentations across age groups. Estimated Resident Population (ERP) data were collected from the Australian Bureau of Statistics for each age group in each year analysed (2018-19 – 2022-23) (ABS, 2024). The latter calendar year was used to calculate the ASR for each age group (i.e., 2019 ERP for 2018-19 counts of dental-related ED presentations). The age-specific rate was calculated by dividing the number of dental-related ED presentations in a specific age group by the total population at risk within that same age group for a given period. Rates are expressed per 100,000 population. Age-specific rates were analysed across all years and according to 79 Victorian Local Government Areas (LGAs). Due to small sample size at the LGA level in the younger and older age groups (0-4, 5-9, 10-19 and 45-64 and 65+), the age groups were aggregated to 0-19 years old and 45+ years old. At the LGA level, counts with low numbers of presentations (<20 each year in each age group) were suppressed due to low sample size (n.p).

Data management and analysis was performed using Microsoft 365 and Stata BE Version 18.0 (StataCorp, 2026).

The study sample was 8,031,725. Episodes were included if the patient had a usual postcode of residence in Victoria and excluded due to incomplete year, missing demographic information and non-emergency visits (including dead on arrival and return visits). Figure 1 illustrates how the study sample was derived.

**Figure 1.**
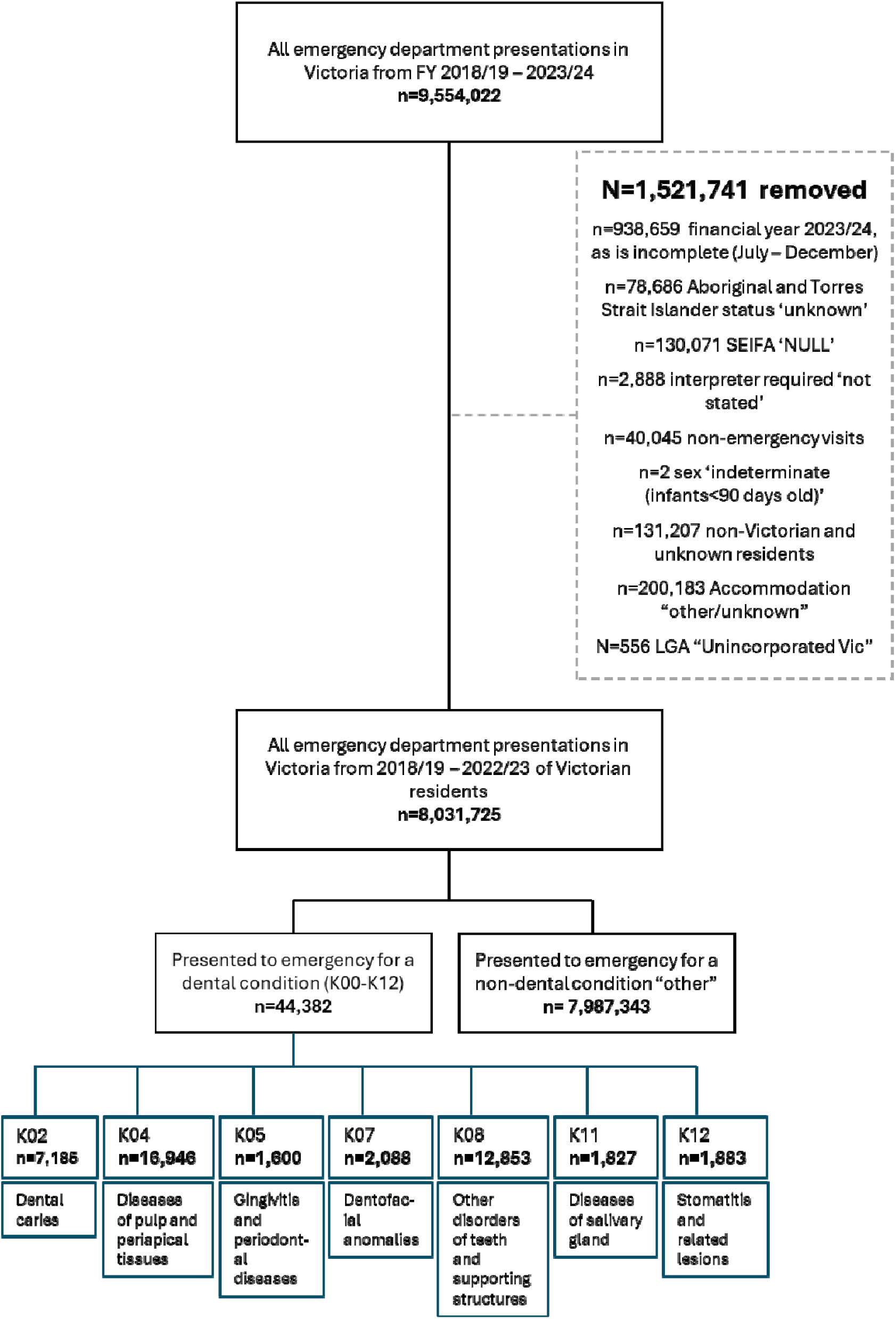
Participant flowchart

## Results

Table 1 presents the demographic profile of Victorian residents presenting to a Victorian ED for a dental-related condition or non-dental condition (Other). Over the years 2018-19 to 2022-23, 0.55% (n=44,382) of ED presentations across Victorian hospitals by residents were dental-related. The slight increase in proportion of dental-related ED presentations in 2018-19 and 2020-21 may be attributed to the Covid-19 pandemic restrictions which limited Victorians access to public oral healthcare for non-emergency or urgent care. Aboriginal Victorians have a presentation rate of 1.10% across all years compared to non-Aboriginal Victorians (0.54%). As the area advantage increases, the proportion of Victorians dental-related ED presentations decreases; 0.72% in quintile 1 (most disadvantage), 0.62% in quintile 2, 0.51% in quintile 3, 0.44% in quintile 4 and 0.38% in quintile 5 (least disadvantage). The proportion of dental-related presentations were higher among populations living in regional and remote areas compared with those in major cities; 0.45% in major cities, 0.77% in inner regional, 0.97% in outer regional and 0.91% in remote areas. People experiencing or at risk of experiencing homelessness (PEH) have a higher proportion of dental-related presentations (0.76%) compared with Victorians in private residences (0.56%) and dependant accommodation (0.16%).

**Table 1.** Characteristics of study population by reason for presenting to ED.

|  | Dental conditions | Other |
| --- | --- | --- |
| Presentations to ED % (n) | 0.55 (44,382) | 99.45 (7,987,343) |
| **Year, % (n)** |  |  |
| 2018-19 | 0.57 (9,100) | 99.43 (1,593,007) |
| 2019-20 | 0.58 (8,977) | 99.42 (1,532,715) |
| 2020-21 | 0.59 (9,136) | 99.41 (1,538,050) |
| 2021-22 | 0.51 (8,279) | 99.49 (1,614,638) |
| 2022-23 | 0.52 (8,890) | 99.48 (1,708,933) |
| **Age, % (n)** |  |  |
| 0-4 | 0.25 (2,225) | 99.75 (885,398) |
| 5-9 | 0.57 (2,183) | 99.43 (378,324) |
| 10-19 | 0.35 (2,669) | 99.65 (760,898) |
| 20-44 | 0.89 (22,963) | 99.11 (2,545,089) |
| 45-64 | 0.66 (10,418) | 99.34 (1,573,524) |
| 65+ | 0.21 (3,924) | 99.79 (1,844,110) |
| **Sex, % (n)** |  |  |
| Male | 0.58 (22,418) | 99.42 (3,865,125) |
| Female | 0.53 (21,951) | 99.47 (4,119,357) |
| Intersex | 0.45 (13) | 99.55 (2,861) |
| **Aboriginal, % (n)** |  |  |
| Yes | 1.10 (1,983) | 98.90 (178,786) |
| No | 0.54 (42,399) | 99.46 (7,808,557) |
| **Preferred Language, % (n)** |  |  |
| English | 0.56 (42,287) | 99.44 (7,486,777) |
| LOTE | 0.42 (2,095) | 99.58 (501,566) |
| **Interpreter Required, % (n)** |  |  |
| Yes | 0.42 (1,301) | 99.58 (305,967) |
| No | 0.56 (43,081) | 99.44 (7,681,376) |
| **IRSD quintiles, % (n)** |  |  |
| Quintile 1 (most disadvantaged) | 0.72 (14,619) | 99.28 (2,003,294) |
| Quintile 2 | 0.62 (10,620) | 99.38 (1,695,448) |
| Quintile 3 | 0.51 (7,305) | 99.49 (1,428,061) |
| Quintile 4 | 0.44 (7,061) | 99.56 (1,598,093) |
| Quintile 5 (least disadvantaged) | 0.38 (4,777) | 99.62 (1,262,447) |
| **Remoteness, % (n)** |  |  |
| Major Cities of Australia | 0.45 (26,070) | 99.55 (5,723,232) |
| Inner Regional Australia | 0.77 (14,300) | 99.23 (1,854,587) |
| Outer Regional Australia | 0.97 (3,974) | 99.03 (405,375) |
| Remote | 0.91 (38) | 99.09 (4,149) |
| **Mental health patient, % (n)** |  |  |
| Yes | 0.05 (54) | 99.44 (103,447) |
| No | 0.56 (44,328) | 99.95 (7,883,896) |
| **Usual Accommodation, % (n)** |  |  |
| Private residence | 0.56 (43,926) | 99.44 (7,753,955) |
| Dependent accommodation | 0.16 (354) | 99.84 (219,998) |
| Homeless | 0.76 (102) | 99.24 (13,400) |

Of the dental-related ED presentations in Victoria, the frequency was highest for those with principal diagnosis code K04 (Diseases of pulp and periapical tissues), K08 (Other disorders of teeth and supporting structures), and K02 (Dental caries) (Figure 2). See Appendix 2 for the number and proportion of dental-related ED presentations by diagnosis type, stratified by year.

**Figure 2.**
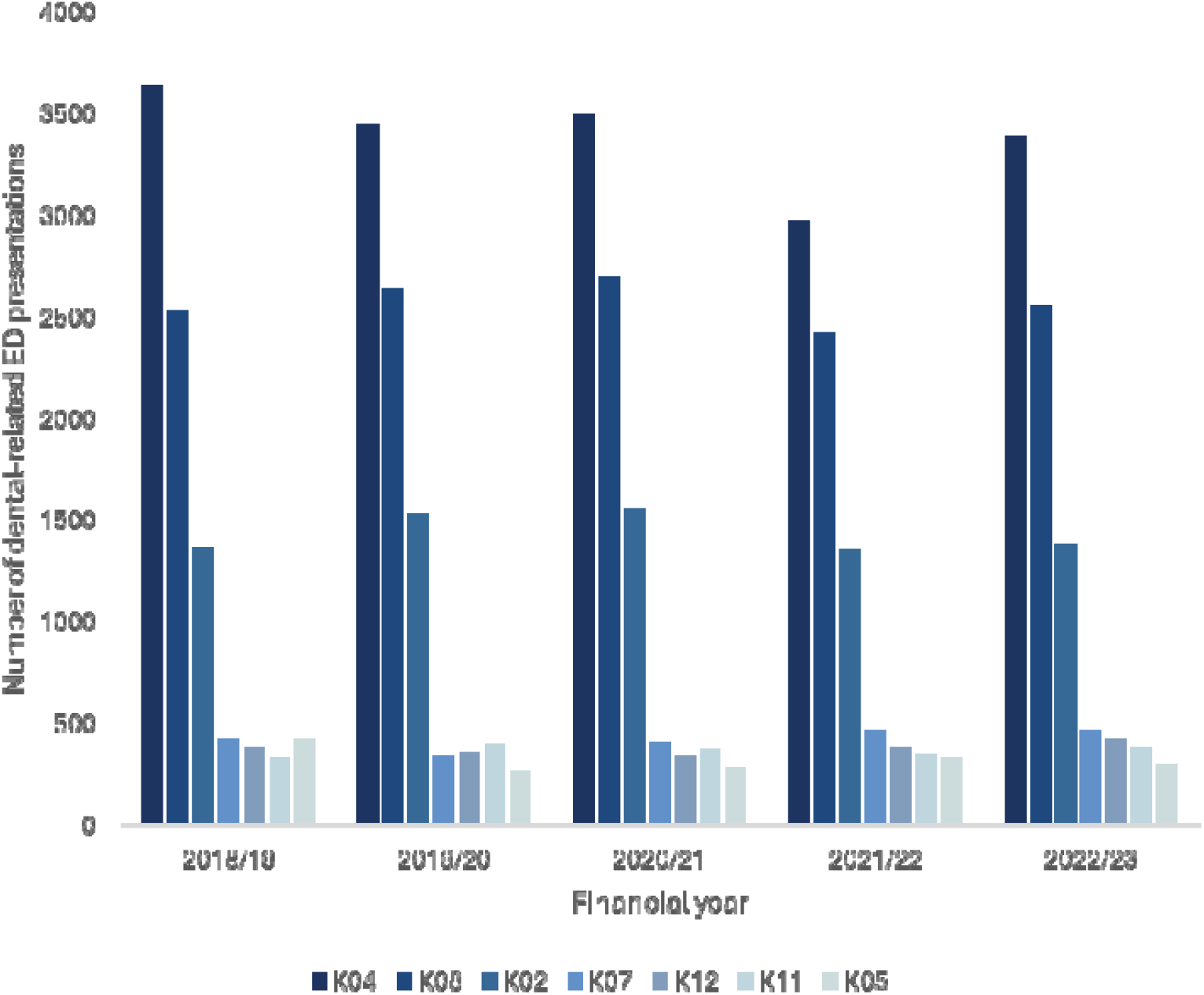
Number of dental-related ED presentations by principal ICD-10-AM diagnosis code, Victorian residents, stratified by financial year.

## Relative risk ratio

Multinomial logistic regression inclusive of all parameter variables spanning 2018-19-2022-23 financial years identified no significant differences for people whose preferred language was not English (see Appendix 1). Consequently, preferred language was excluded from the final model fit (Table 2).

**Table 2.** The association between patient characteristics and dental-related ED presentations combined years 2018-19 – 2022-23. Model 2= Multinomial logistic regression of all parameters excluding preferred language.

| 2018-19-2022-23 | Model 2 | p-value |
| --- | --- | --- |
| Age group<br>0-4 years | Ref | <0.001 |
| 5-9 years | 2.26 (2.13,2.40) |  |
| 10-19 years | 1.35 (1.28,1.43) |  |
| 20-44 years | 3.61 (3.46,3.77) |  |
| 45-64 years | 2.57 (2.45,2.69) |  |
| 65+ years | 0.85 (0.81,0.90) |  |
| **Sex** |  |  |
| Female | Ref |  |
| Intersex | 0.82 (0.47, 1.41) | 0.469 |
| Male | 1.17 (1.15, 1.19) | <0.001 |
| **Aboriginal** |  |  |
| No | Ref |  |
| Yes | 1.49 (1.42,1.56) | <0.001 |
| **Interpreter required** |  |  |
| No | Ref |  |
| Yes | 1.13 (1.07, 1.20) | <0.001 |
| **Remoteness** |  |  |
| Major city | Ref |  |
| Inner regional | 1.60 (1.56,1.63) | <0.001 |
| Outer regional | 1.92 (1.85,1.99) |  |
| Remote | 1.79 (1.30,2.46) |  |
| **IRSD** |  |  |
| Quintile 1 | Ref |  |
| Quintile 2 | 0.92 (0.90,0.94) | <0.001 |
| Quintile 3 | 0.81 (0.79,0.83) |  |
| Quintile 4 | 0.74 (0.72,0.77) |  |
| Quintile 5 | 0.67 (0.65,0.69) |  |
| **Accommodation** |  |  |
| Private Residence | Ref |  |
| Dependent | 0.47 (0.43,0.53) | <0.001 |
| Homeless | 0.92 (0.75,1.12) | 0.383 |
| *Model 1 which includes preferred language can be found in Appendix 1* |  |  |

Across all years, 20-44 year olds have the highest RRR compared to 0 – 4-year-olds (reference) being 3.61 (95%CI: 3.45, 3.77, p<0.001) times as likely to present to ED for a dental-related condition, followed by 45 – 64-year-olds (2.57, 95%CI:2.45, 2.69, p<0.001) and 5 – 9-year-olds (2.26, 95%CI: 2.13, 2.40, p<0.001). Males and females have similar RRR (1.17, 95%CI: 1.15, 1.19, p<0.001). Aboriginal Victorians have a higher RRR (1.49, 95%CI: 1.42, 1.56, p<0.001) compared to non-Aboriginal Victorians. Requiring an interpreter is associated with a 1.13-time increased relative risk (RR) (95%CI: 1.10, 1.31, p<0.001) for dental-related ED presentations. Outer regional areas have the greatest increase in RRR compared to major cities (1.91, 95%CI: 1.85, 1.99, p<0.001), followed by remote (1.79, 95%CI: 1.30, 2.46, p<0.001). There is an inverse relationship with the IRSD and dental-related ED presentations, as socio-economic advantage increases, the RR of presenting to ED for a dental condition decrease. Compared to quintile 1 (most disadvantage), populations from quintile 2, 3, 4 and 5 have a RRR of 0.92 (95%CI: 0.90, 0.94, p<0.001), 0.81 (95%CI: 0.79, 0.83, p<0.001), 0.74 (95%CI: 0.72, 0.77, p<0.001) and 0.67 (95%CI: 0.65, 0.69, p<0.001). Compared to Victorians living in a private residence, PEH have a RRR of 0.92 (95%CI: 0.75,1.12, p<0.001), and Victorians living in dependent accommodation have a RRR of 0.47 (95%CI: 0.43, 0.53, p=0.383).

Table 3 presents the RRR of the association between patient characteristics and dental-related ED presentations stratified by year. Across the years, 20 – 44-year-olds have the greatest increase in RR compared to 0 – 4-year-olds for dental-related ED presentations. Across the years, identifying as Aboriginal was associated with an increased RRR of dental-related ED presentations compared to Victorians who do not identify as Aboriginal. Across all years, the RRR of dental-related ED presentations decreases as socio-economic advantage increases.

**Table 3.**
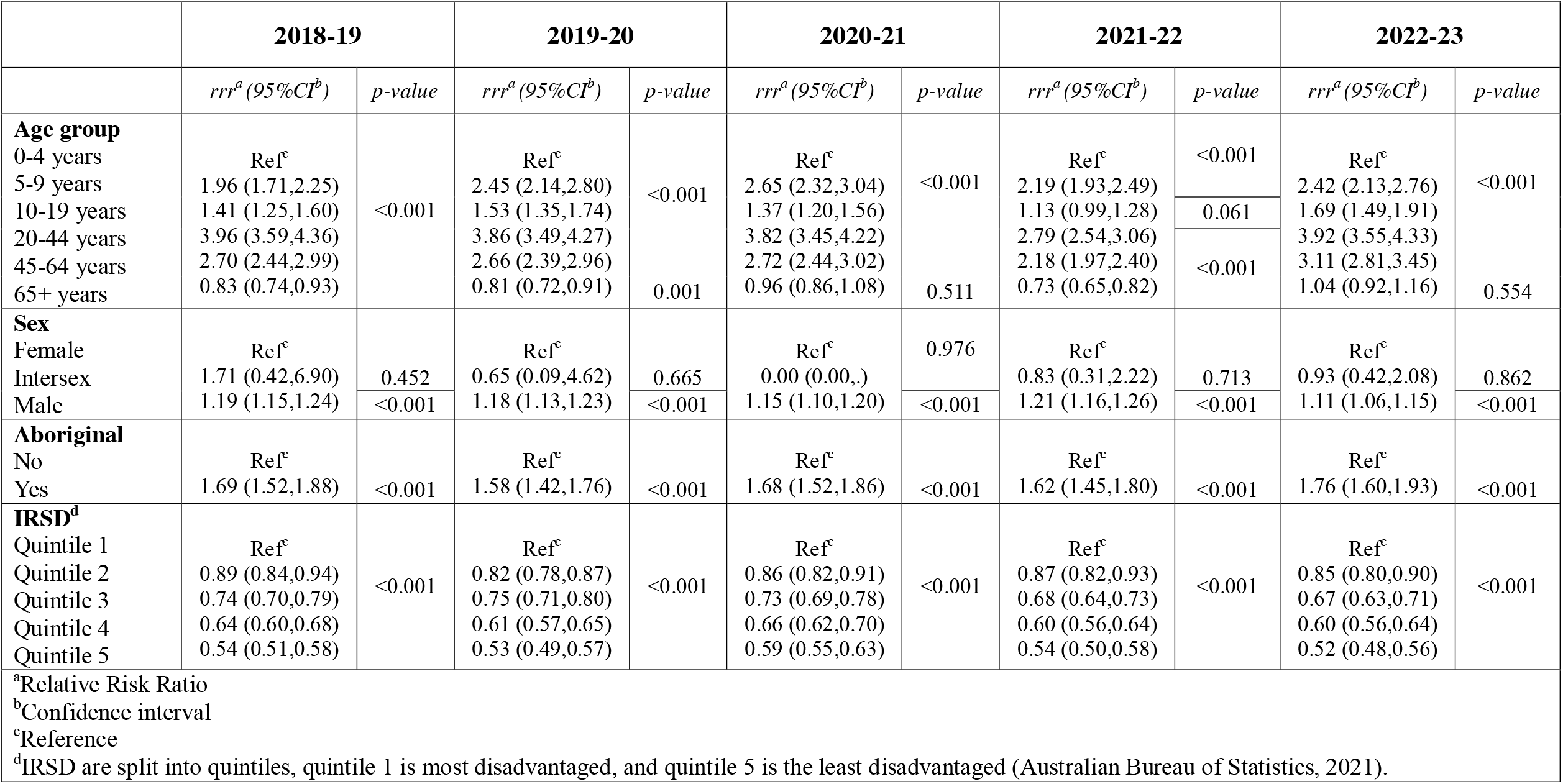
The association between patient characteristics and presenting to a Victorian emergency department for a dental condition stratified by year.

**Table 4.**
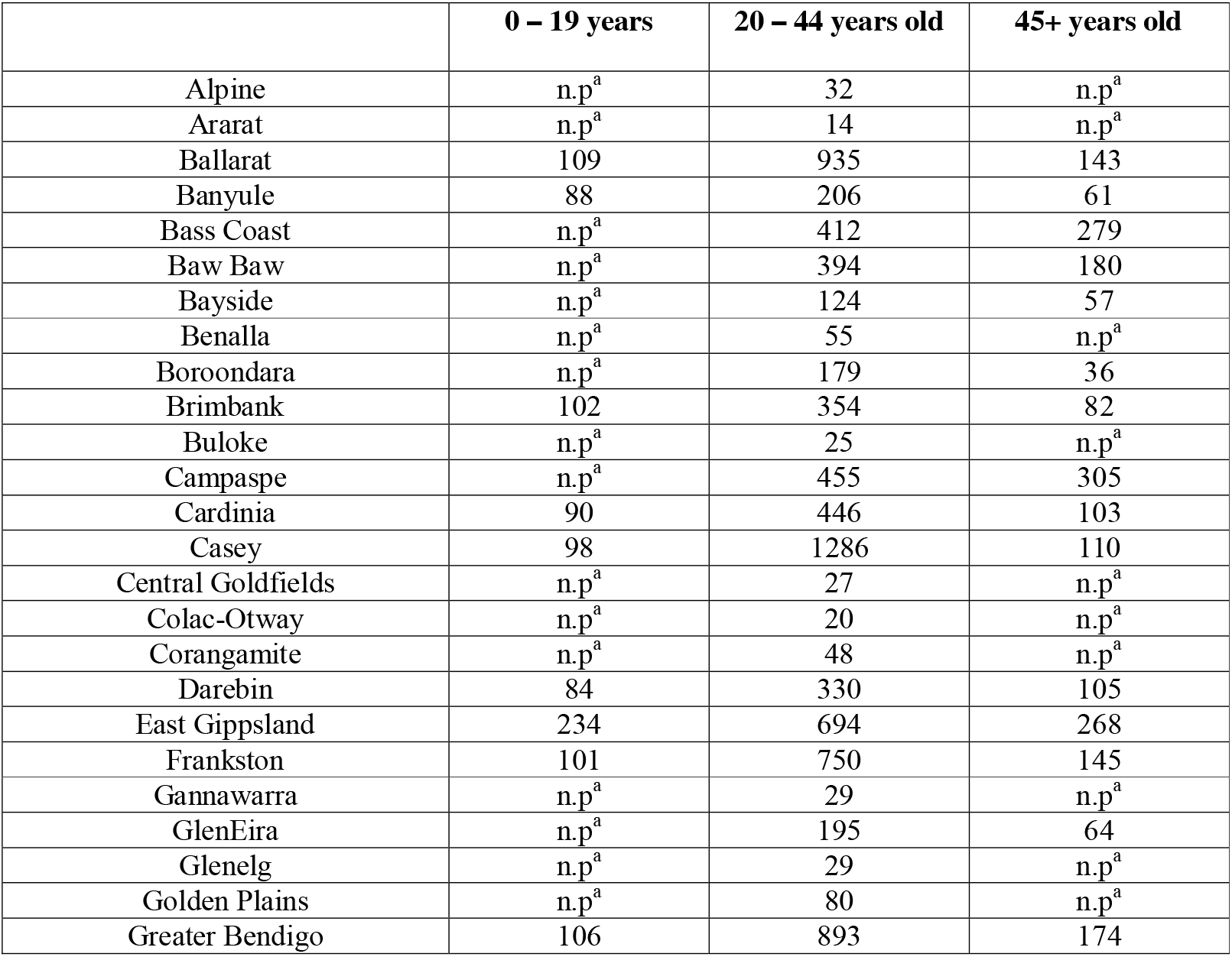
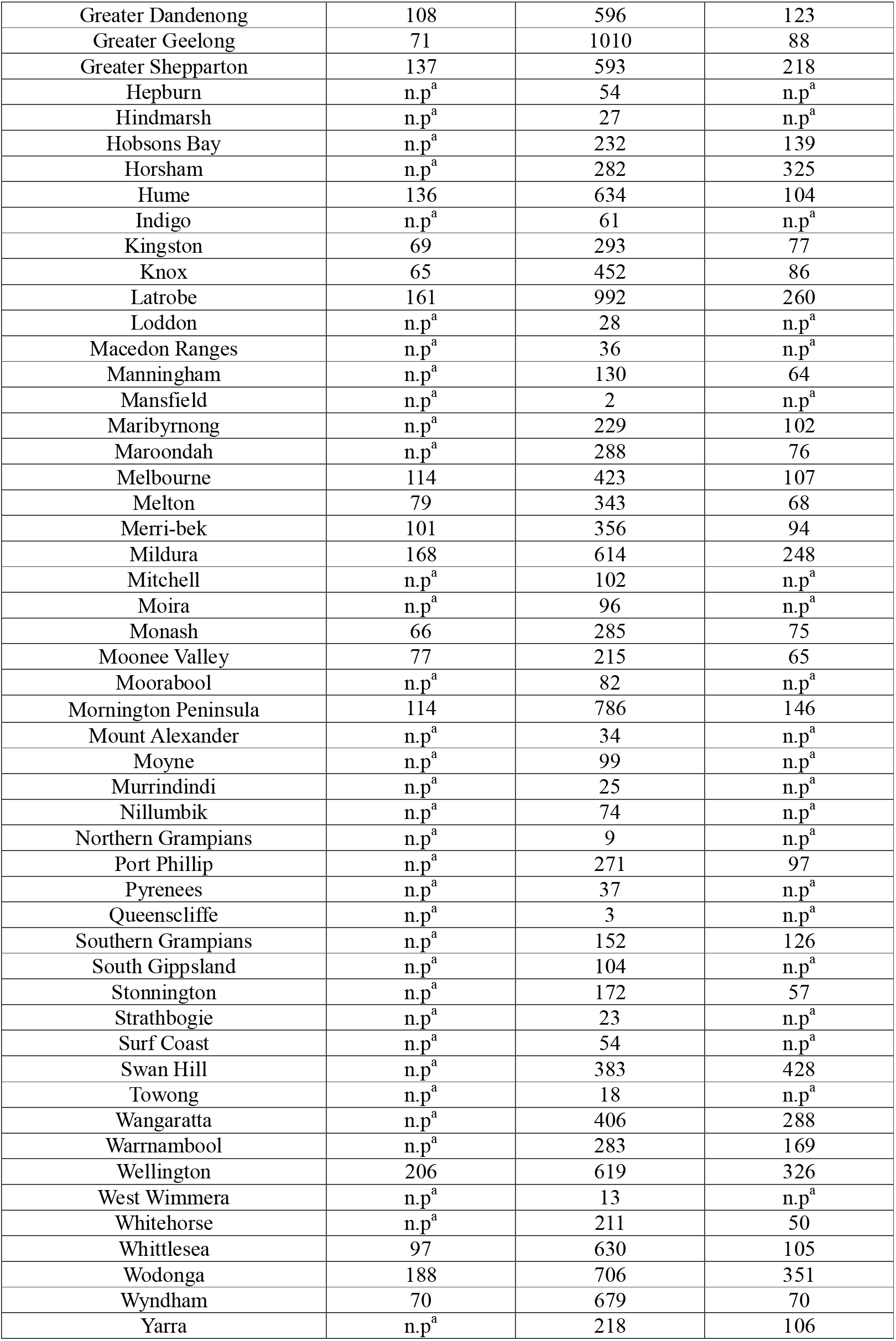
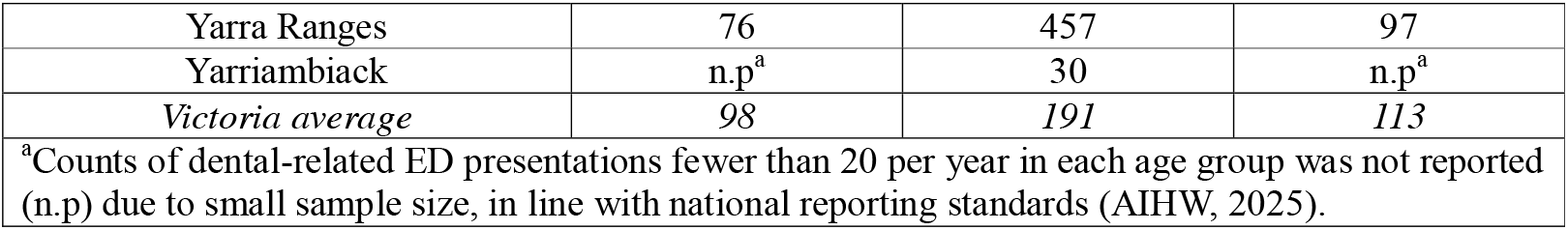
Age-specific rates per 100,000 population of ED dental-related presentations stratified by age group and LGA across all years.

## Age-specific rates

Among LGAs, East Gippsland had the highest age-specific rates across both the 0 – 19 and 20 – 44 age groups at 234 and 1,280 per 100,000 population, respectively across all years. In the 45+ age group, Swan Hill had the highest rate across all years at 428 per 100,000 population. 10 LGAs had higher than overall Victorian age-specific rates of dental-related ED presentation across all age groups and years: Ballarat, East Gippsland, Frankston, Greater Bendigo, Greater Shepparton, Latrobe, Mildura, Mornington Peninsula, Wellington, and Wodonga.

## Discussion

The present study findings indicate low dental-related ED presentations amongst Victorians at Victorian ED (0.55%) relative to other states of Australia (0.91%-1%) (Musa et al., 2025; Verma & Chambers, 2014). Over the study period, the proportion of Victorians presenting to ED for dental conditions remained relatively stable with a slight increase in 2018-19 and 2020-21. Covid-19 was first identified in Australia in January of 2020. By March 2020, Victoria had declared a state of emergency (Premier of Victoria, 2020) which came with movement restrictions and limited access to oral healthcare services with the suspension of non-urgent oral health care for most of 2020 and into 2021 (Minister for Health, 2020; Nguyen et al., 2021).

There is a statistically significant association between Aboriginal Victorians and dental-related ED presentations compared to non-Aboriginal Victorians. Similarly, socio-economic status was significantly associated with dental-related ED presentations. The present study found that living in areas of greater socio-economic disadvantage increased the likelihood of dental-related ED presentations, which is consistent with existing literature on the social determinants of oral health which document the greater burden of oral disease amongst individuals from lower socio-economic status (de Abreu et al., 2021). This finding may support international research by Obeidat and colleagues on the impact of social determinants on oral health, who found that people of higher socio-economic status are more likely to have better oral health seeking behaviours (regular dental visits) and perceive their oral health positively (Obeidat et al., 2024).

Compared to Victorians living in a private residence, PEH and Victorians living in dependent accommodation are less likely to present to ED for dental conditions. Research shows that PEH generally seek care through ED due to barriers to accessing primary care, including oral healthcare (Durey et al., 2022; Miller et al., 2024; Osman et al., 2026). Hence, reduced dental-related ED presentation rates among PEH in the present study is unlikely to infer improved oral health outcomes, but rather, that PEH are less likely to present to ED for dental conditions, compared to other health conditions. This aligns with existing literature exploring PEH presentation to ED in Victorian ED being primarily for mental health, behavioural/intoxication, soft tissue injury/infection and abdominal complaints (Osman et al., 2026). In Victoria, PEH are eligible for public oral healthcare, and are given priority access to general oral healthcare through the Priority Access policy (Oral Health Victoria). This provides PEH with the next available appointment and allows them to bypass the waiting lists. However, due to non-economic barriers to care, stigma and the complex medical and psychosocial health needs of PEH, oral health is often disregarded (Paisi et al., 2020).

Although oral healthcare maybe more costly for PEH compared to traditional models of care, research has demonstrated cost-effective examples of outreach services for PEH (Nguyen et al., 2024).

Amongst the 10 LGAs with age-specific rates of dental-related ED presentations higher than state-wide levels, most are from areas of greater socio-economic disadvantage. Greater Shepparton, Latrobe, Mildura are from Quintile 1 on IRSD. Ballarat, East Gippsland, Greater Bendigo, Wellington and Wodonga are from Quintile 2. Frankston is from Quintile 3 and Mornington Peninsula is from Quintile 4. All LGAs except Frankston are in regional Victoria. This present study reinforces findings from previously published literature about the association between socio-economic disadvantage and worse oral disease (Singh et al., 2025). Whilst the findings of Singh and colleagues were limited to the public oral healthcare sector; our present study looked at the wider Victorian population and identified similar trends and associations, with higher rates of presentations and increased RRR of presentation in people from areas with greater socio-economic disadvantage.

### Limitations and opportunities for further research

This analysis was limited to Victorians that presented to Victorian EDs, to understand the local trends in utilisation of EDs for dental conditions. Previous studies in other Australian states and territories have explored the outcome of interest for their constituents and found similar results (Musa et al., 2025; Verma & Chambers, 2014). Further research across the country exploring the differences in the rates and associated demographic factors would be beneficial to identify variations at the state and territory level. Furthermore, additional data on individual factors such as concession card holder status, private health insurance status, and risk factors such as sugar sweetened beverage consumption and oral hygiene behaviours would be useful in identifying the impact of individual level health and behavioural factors to help mitigate ED presentations for dental conditions.

Dental-related ED presentations are a strain on already burdened hospitals across Victoria; Medicare Urgent Care Clinics provide healthcare to people who need urgent care but not an emergency response, offer no cost, walk-in access to GPs without a referral (Lucy MacDonald, 2023). Currently, there are 38 locations across Victoria. Additional awareness of these avenues for access to timely healthcare outside of the hospital, for non-life-threatening health, including, dental-related conditions, may be useful in reducing the burden on the public healthcare system in Australia. However, additional training and clinical support may be needed for health practitioners working in Medicare Urgent Care Clinics, given competencies in oral health assessment and management are often excluded within the education training of non-dental health professionals (Smith et al., 2024). The VVED provides Victorians with access to an ED professional from their home. While this current study included presentations to VVED, further analysis into presentations that were through the VVED compared to physical presentations was not examined. Oral health training for VVED staff would ensure Victorians have equitable access to oral health information when presenting to VVED for a dental-related condition.

## Conclusion

The frequency of ED presentations for dental conditions can place a significant strain on the public healthcare system in Victoria, diverting healthcare resources from where they may be needed most. This study has explored the frequency, associations and rates of Victorians presenting to Victorian ED for dental conditions and revealed that patient demographic factors, especially socio-economic status, may be useful in identifying the relative risk of individuals for dental-related ED presentations. There is an opportunity for further research to explore the impact of individual health and behavioural factors on Victorians’ presenting to ED for dental conditions that are preventable, and the influence of non-hospital urgent care services in reducing the burden on ED in Victoria.

## Data Availability

All data produced in the present study are available upon reasonable request to the authors, subject to data custodian approval.

## Acknowledgements

We would like to acknowledge Aaron Bloch for his contributions.

## Ethics approval statement

This study received ethics approval by Deakin University Human Research Ethics Committee (DUHREC 2024-054).

