## Appendix 1 for "An investigation of dental-related emergency department presentations and patient characteristics in Victoria, Australia"

### **Appendix 1.** Model 1: Multinomial logistic regression of all parameter variables

| **2018/19-2022/23** | **Model 1** | **p-value** |
| --- | --- | --- |
| **Age group**  0-4 years  5-9 years  10-19 years  20-44 years  45-64 years  65+ years | Ref  2.26 (2.13, 2.40)  1.35 (1.28, 1.43)  3.61 (3.46, 3.77)  2.57 (2.45, 2.69)  0.85 (0.81, 0.90) | <0.001 |
| **Sex**  Female  Intersex  Male | Ref  0.82 (0.47, 1.41)  1.17 (1.15, 1.19) | 0.466  <0.001 |
| **Aboriginal**  No  Yes | Ref  1.49 (1.42, 1.56) | <0.001 |
| **Preferred language other than English**  No  Yes | Ref  0.94 (0.88, 1.01) | 0.097 |
| **Interpreter required**  No  Yes | Ref  1.20 (1.10, 1.31) | <0.001 |
| **Remoteness**  Major city  Inner regional  Outer regional  Remote | Ref  1.60 (1.56, 1.63)  1.91 (1.85, 1.98)  1.78 (1.29, 2.46) | <0.001 |
| **IRSD**  Quintile 1  Quintile 2  Quintile 3  Quintile 4  Quintile 5 | Ref  0.92 (0.90, 0.94)  0.81 (0.79, 0.83)  0.74 (0.72, 0.76)  0.67(0.65, 0.69) | <0.001 |
| **Accommodation**  Private Residence  Dependent  Homeless | Ref  0.47 (0.43, 0.53)  0.92 (0.75, 1.11) | <0.001  0.383 |
