## Appendix 2 for "An investigation of dental-related emergency department presentations and patient characteristics in Victoria, Australia"

### **Appendix 2.** Number and proportion of dental-related ED presentations, by diagnosis type stratified by year

|  | **K02^a^** | **K04^a^** | **K05^a^** | **K07^a^** | **K08^a^** | **K11^a^** | **K12^a^** | **Other^b^** | **Total** |
| --- | --- | --- | --- | --- | --- | --- | --- | --- | --- |
| **2018/19, n (%)** | 1,364 (0.1 %) | 3,648 (0.2 %) | 427 (0.0 %) | 427 (0.0 %) | 2,531 (0.2 %) | 326 (0.0 %) | 380 (0.0 %) | 1,593,128  (99.4 %) | **1,602,231**  **(100.0 %)** |
| **2019/20, n (%)** | 1,532 (0.1 %) | 3,445 (0.2 %) | 267 (0.0 %) | 335 (0.0 %) | 2,641 (0.2 %) | 398 (0.0 %) | 359 (0.0 %) | 1,532,819  (99.4 %) | **1,541,796**  **(100.0 %)** |
| **2020/21, n (%)** | 1,556 (0.1 %) | 3,493 (0.2 %) | 281 (0.0 %) | 405 (0.0 %) | 2,696 (0.2 %) | 371 (0.0 %) | 334 (0.0 %) | 1,538,170  (99.5 %) | **1,547,306**  **(100.0 %)** |
| **2021/22, n (%)** | 1,355 (0.1 %) | 2,974 (0.2 %) | 327 (0.0 %) | 463 (0.0 %) | 2,426 (0.2 %) | 351 (0.0 %) | 383 (0.0 %) | 1,614,753  (99.5 %) | **1,623,032**  **(100.0 %)** |
| **2022/23, n (%)** | 1,355 (0.1 %) | 2,974 (0.2 %) | 327 (0.0 %) | 463 (0.0 %) | 2,426 (0.2 %) | 351 (0.0 %) | 383 (0.0 %) | 1,614,753  (99.5 %) | **1,623,032**  **(100.0 %)** |
| **Total, n (%)** | 1,378 (0.1 %) | 3,388 (0.2 %) | 298 (0.0 %) | 458 (0.0 %) | 2,560 (0.2 %) | 381 (0.0 %) | 427 (0.0 %) | 1,709,026  (99.5 %) | **1,717,916**  **(100.0 %)** |
| ^a^Diagnosis types**:** K02- Dental caries, K04- Diseases of pulp and periapical tissues, K05- Gingivitis and periodontal diseases, K07- Dentofacial anomalies K08- Other disorders of teeth and supporting structures, K11- Diseases of salivary glands and K12- Stomatitis and related lesions  ^b^Non-dental conditions | | | | | | | | | |
